# Deciphering the cellular tumor microenvironment landscape in salivary gland carcinomas using multiplexed imaging mass cytometry

**DOI:** 10.1101/2025.05.11.25327400

**Authors:** Marcel Mayer, Lisa Nachtsheim, Louis Jansen, Philipp Wolber, Marcel Schmiel, Alexander Quaas, Jens Peter Klussmann, Christoph Arolt

## Abstract

**Purpose:** To spatially characterize the single-cell tumor microenvironment (TME) of salivary gland carcinomas (SGC) and identify prognostic biomarkers.

**Experimental Design:** SGC, including salivary duct carcinomas (SDC), acinic cell, mucoepidermoid, and secretory carcinomas, were analyzed using a 13-marker imaging mass cytometry panel. Multichannel image data from 54 primary cases and nodal metastases were processed to generate single-cell datasets. Cell phenotypes (tumor cells, cancer-associated fibroblasts (CAFs), endothelia, immune cells) were classified using a validated CAF algorithm, followed by spatial analysis and clinicopathological correlation.

**Results:** Among 509,364 cells, SDC exhibited the highest fractions of Collagen-and matrix-CAFs (mCAFs). Acinic cell carcinomas (ACC) showed enriched CD4+/CD8+ T cells and antigen-presenting CAFs (apCAFs), indicating strong immune infiltration. A spatially defined cellular neighborhood (CN8) of mCAFs and endothelia was elevated in SDC, with higher CAF infiltration in androgen receptor (AR)_high_ versus AR_low_ SDC. Elevated mCAF frequency and CN8 were significantly associated with reduced recurrence-free probability (RFP) and distant control rates (DCR). Additionally, higher mCAF frequencies were an independent prognostic factor for decreased RFP and DCR in Cox regression analysis.

**Conclusion:** SDC are characterized by Collagen-/mCAF-rich microenvironments and mCAF-endothelial spatial interactions that are linked to metastasis. ACC display pronounced immune infiltration, suggesting its potential for immunotherapy. mCAFs in SDC emerge as prognostic biomarkers and therapeutic targets, highlighting the importance of targeting CAF-driven metastasis in future treatments. This study provides insights into the biology of SGC and identifies novel prognostic markers.

## Introduction

Salivary gland carcinomas (SGC) are rare and account for approximately 6% of head and neck malignancies (1). They form a heterogeneous group of 21 morphologically and prognostically distinct tumor entities (2), some of which have high potential for recurrence and metastasis. Apart from biomarker-guided therapies for salivary duct carcinomas (SDC) and secretory carcinomas (Sec), there is currently no established targeted systemic therapy for SGC.

The tumor microenvironment (TME), which consists of a cellular compartment and the extracellular matrix (ECM), is an essential component of carcinomas and significantly influences their growth and response to chemotherapy (3). Cancer-associated fibroblasts (CAFs) are activated fibroblasts that form the cellular TME, together with immune cells and tumor vasculature. Previous studies have characterized a variety of CAF subpopulations, each with a distinct marker profile, functionality, and spatial distribution (4–7). However, because of the requirement for several markers, the precise characterization of CAF subsets remains a challenge when analyzing larger patient cohorts. Various studies have examined the importance of CAFs in carcinogenesis, ECM synthesis, angiogenesis, immune modulation, and chemotherapy resistance (8–10). As anticipated, CAFs also influence patient outcome (11, 12). Pro-carcinogenic factors make CAFs promising targets for new therapeutic approaches (13). Cords et al. recently published two studies that systematically characterized CAF subsets across carcinomas from different organs using an unbiased single-cell RNA sequencing (scRNA-seq) approach and proposed a generalized CAF classification concept. They methodically transferred their classification algorithm to space-agnostic imaging mass cytometry (IMC) and demonstrated that CAF subtypes reside in distinct spatial niches, partly correlate with reduced immune infiltrate, and can determine prognosis in non-small cell lung cancer (NSCLC) (5, 12). To date, little is known about the occurrence and function of CAFs in SGC. In a pilot study, we demonstrated significant differences in the frequency of COL11A1+ CAFs between different SGC entities (14). More recently, we analyzed a large SGC cohort using deep proteomic profiling and demonstrated that SGC can be subclassified into two large groups based on the ECM, which, among other parameters, differentiates with regard to immune cell infiltration (15). Given the marked impact of CAF subtypes on patient outcomes and the potential of TME-directed therapies, we aimed to analyze the cellular TME of 54 patients with SGC. This included four different tumor entities without myoepithelial differentiation, as myoepithelial cells may show an overlapping immunophenotype with CAFs when using a restricted marker panel. Using a multiplex IMC panel and a thoroughly validated classification scheme, we analyzed the spatial distribution of CAF subtypes, immune cells, tumor vasculature, and tumor cells, and correlated our findings with clinical data and the ECM landscape to shed light on the cellular TME landscape of SGC.

## Methods

### Patient cohort

The cohort comprised patients with sufficient formalin-fixed paraffin-embedded (FFPE) material of the primary tumor and/or lymph node metastasis collected at first diagnosis who were treated for primary SGC at the Department of Otorhinolaryngology, Head and Neck Surgery of the University Hospital of Cologne, Germany between January 1^st^, 2004, and December 31^st^, 2021. Demographics, survival, and histopathological data were extracted from clinical records and histopathological reports with respect to entities and disease stage at the time of diagnosis according to the AJCC TNM staging system (8th edition, 2020) (16). In cases of missing follow-up data within the clinical records, patients and/or their general practitioners were phoned to follow up on the current tumor status. The study was conducted in accordance with the Declaration of Helsinki and was approved by the Ethics Committee of the University of Cologne (Approval Code: 13-091). Informed consent was obtained from all participants included in this study. Because this was not a prospective clinical trial, formal power calculations and randomization were not performed.

### Antibody panel and conjugation of antibodies with metal isotopes

A panel including 15 markers that distinguish CAFs, immune cells, tumor cells, and endothelia was designed. The following monoclonal antibodies were purchased from Fluidigm^®^: CKAE1/3-148Nd (dilution 1:100; RRID:AB_3678170), CD31-146Nd (dilution 1:200; RRID:AB_3678176), Vimentin-143Nd (dilution 1:50; RRID:AB_2811069), aSMA-141Pr (dilution 1:200; RRID:AB_2890139), Collagen1-173Yb (dilution: 1:300; RRID:AB_3678205), Ki67-168Er (dilution: 1:50; RRID:AB_2800467), CD34-151Eu (dilution 1:200; RRID:AB_3678086), CD74-166Er (1:50; RRID:AB_3675795), CD45-152Sm (dilution: 1:50; RRID:AB_3678248), CD4-156Gd (dilution: 1:100; RRID:AB_2811051), CD8a-162Dy (dilution 1:200; RRID:AB_2811053), IDO-155Gd (dilution: 1:100; RRID:AB_3678089), and CD73-158Gd (dilution: 1:100; RRID:AB_3678093). The carrier-free CD138 monoclonal antibody (mouse, clone: B-A38; RRID:AB_3676290) was purchased from Origene^®^ and conjugated with 164Dy (dilution 1:100) using the Maxpar X8 Multimetal Labeling Kit (Fluidigm^®^, 201300), following the manufacturer’s protocol. The intercalator iridium (Fluidigm^®^, 2011922 B) was included for the identification of nuclei. The conjugated antibodies were titrated and tested for specificity by visual comparisons in SGC samples, as well as in ovarian, breast, lung, and colon carcinoma control samples, resulting in the abovementioned dilutions for optimal signal yield.

### Staining

The tissue was freshly sectioned and placed onto tissue microarray (TMA) slides five days before staining. Tissue slides underwent imaging mass cytometry (IMC) staining following the Fluidigm^®^ protocol (PN400322A3). FFPE tumor samples were baked at 65°C for two hours to remove residual wax. First, deparaffinization was performed. The slides were rehydrated in an ethanol series (100%, 95%, 90%, 80%, and 70%). Antigen retrieval was performed in a water bath at 96°C using Tris-EDTA buffer (pH 9) for 30 minutes. Next, the slides were blocked with 3% bovine serum albumin (BSA) in Maxpar phosphate-buffered saline (PBS) for 45 minutes at room temperature in a hydration chamber. Afterwards, the slides were incubated with the antibody cocktail in the above-mentioned dilutions (0.5% BSA buffer) and stored overnight at 4°C in a hydration chamber. The following day, the slides were washed four times with 0.2% Triton X-100 in Maxpar PBS and twice with Maxpar PBS. DNA staining was performed by incubating the slides with Intercalator-Ir (Fluidigm^®^, 201192 B) in Maxpar PBS (dilution 1:400) for 30 min in a hydration chamber. Finally, the slides were washed twice with Maxpar water and air-dried for 30 minutes.

### Imaging mass cytometry

Over a period of four weeks, all cores on all six TMA slides were laser-ablated at 200 Hz with an ablation energy of 3 dB using a Hyperion imaging mass cytometry system connected to a Helios mass cytometer (Fluidigm^®^). A tuning slide (Fluidigm^®^) was used prior to each TMA slide. All ROIs were set to 500 μm × 500 μm.

### Preprocessing

The IMC raw data were preprocessed and segmented using a combination of Jupyter notebooks (RRID:SCR_018315), CellProfiler pipelines (RRID:SCR_007358), and the Ilastik tool (RRID:SCR_015246) as described previously (12, 17). Briefly, using published Jupyter notebooks (RRID:SCR_018315), proprietary MCD files were converted into single-channel TIFFs, filtered for “hot pixels,” upscaled, and cropped to facilitate the segmentation step. Then, an Ilastik pixel classifier was trained, which used all marker channels except Ki67. Because CAFs can have extensive cellular processes, cell bodies were reduced to their nuclear core to enable segmentation of individual cells (12). Next, a CellProfiler pipeline (RRID:SCR_007358) was used to extract single-cell information as a.csv file. All subsequent steps were performed in R using the imcRtools package (RRID:SCR_017132) and published workflow (17). After reading the data in R, the counts were transformed using an inverse hyperbolic sine function. This was followed by optical and numerical verification of the segmentation quality. T-SNE and UMAP plots revealed no batch effect among non-neoplastic cells that could have been introduced by TMA slides (Supplementary Figure S1A and B). All TMA cores were visualized and those without tumor cells were excluded from subsequent analyses. This led to the exclusion of 58 cores (100,425 cells) and a remainder of 199 cores (408,939 cells). These cores from 54 patients comprised 188 and 11 cores from primaries and metastases, respectively.

### Phenotyping

First, the tumor cells were identified using a Gaussian mixture model of cytokeratin expression (Cytokeratin AE1/3 antibody). Next, all ROIs were plotted with tumor cell annotations to ensure morphologically valid classification. All subsequent stepwise clustering of TME cells was performed using a combination of FlowSOM (RRID:SCR_016899) clustering and meta-clustering with ConsensusClusterPlus (RRID:SCR_016954), as described previously (5). First, all TME cells were clustered into endothelial cells (CD31+, Cytokeratin-), immune cells (CD45+/CD138+, Cytokeratin-), and CAFs (CD31-, Cytokeratin-, CD45-, CD138-). Negative selection of CAFs has been previously carried out in other exploratory studies (18). Immune cells were then clustered by the expression of CD45, CD4, CD8, and CD138 into plasma cells (CD138+), CD8+ T cells, CD4+ T cells (CD4^high^), and other CD4-expressing immune cells (CD45+, CD4^low^). The latter cell population likely corresponds to other CD4-expressing hematopoietic cells, such as macrophages and other myeloid-derived cells, which have been shown to express CD4 to a lesser extent using IMC (5, 12, 19). A more detailed subclassification was not feasible based on the choice of markers. CD4+ T cells were clustered again, using CD45, CD4, CD74, and Ki67, into CD4+ T cells, proliferating CD4+ T cells (Ki67^high^), and CD4+CD74+ T cells. This CD4+ T cell subset has recently been found to correspond mostly to tumor-associated Tregs and more specifically to Tregs with effector cytokine production (20, 21).

Next, CAFs were subclassified by clustering with the markers Vimentin, aSMA, CD34, IDO, CD73, CD74, Ki67, and Collagen1 into Collagen CAFs, matrix CAFs (mCAF), SMA+ CAFs (SMA CAF), developing CAFs (dCAF) and antigen-presenting CAFs (apCAF), largely following a classification scheme recently proposed and validated in different carcinoma types (5, 12). Cells without specific CAF marker expression (SMA/Collagen1/Ki67/CD74) were split into CAFs with high Vimentin expression and other cells. The latter clustered again and were shown to correspond to other specific CAF and immune cell subsets. The remaining cells that were negative for all markers in the panel were designated as tumor cells because they were located in tumor cell patches. We reasoned that these cells likely express low levels of cytokeratin which could not be detected with the chosen dilution of the anti-cytokeratin antibody after preliminary experiments with a more limited number of tumor samples. Moreover, since the main scope of this work was the TME, this conservative approach minimizes the danger of producing false-positive results.

### Spatial analysis

Because we assumed different spatial contexts in metastases and primaries, we included only primaries in the subsequent spatial analyses with *imcRtools* (RRID:SCR_017132). First, we constructed a cellular interaction graph using k-nearest neighbor detection with k = 20. We then defined the cellular fractions of the 20 nearest neighbors of each cell and clustered each cell based on this information into one of nine clusters, that is, cellular neighborhoods. We also calculated the results for 6 and 12 clusters but found that nine neighborhoods yielded interesting spatial cellular distributions, such as a tumor-stroma interface neighborhood, without splitting the data into clusters with only one cell type. We then carried out tumor patch detection (minimal patch size of 10 cells) to allow for the identification of tumor-infiltrating cells and the calculation of the spatial distances between each TME cell and the tumor border. Finally, we carried out statistical testing of interactions/co-localization by calculating the averaged spatial interaction of each combination of the two cell types and comparing it against a null distribution in each ROI using the *testInteractions* function from the *imcRtools* package (RRID:SCR_017132), leveraging a testing strategy proposed by Schapiro et al. (22).

### Correlation with SGC ECM modules

We recently proposed three different ECM protein modules, which explain the ECM of SGC and are heavily enriched in CAF-associated terms, basement membrane-associated terms, and those that are linked to peripheral blood components as well as blood-producing and blood-processing organs (15). We found that these modules describe ECM differences between SGC with and without myoepithelial differentiation. From the single-cell dataset, we calculated the median cell type frequency per primary and correlated these values with the ECM module *Eigengenes* in 40 samples that were present in both cohorts using the Spearman method. *P-values* were corrected for multiple testing using the Benjamini-Hochberg procedure.

### Clinico-pathological analysis

Statistical analyses were performed using R software (version 4.4.1) in RStudio (version 2024.4.2.764; RRID:SCR_000432). The R packages *ggplot2* (RRID:SCR_014601)*, survfit, survminer* (RRID:SCR_021094), *survival* (RRID:SCR_021137), *forestmodel* (RRID:SCR_025250), *haven, broom* (RRID:SCR_026874), *lazyeval, tidyverse* (RRID:SCR_019186), *rstatix* (RRID:SCR_021240), and *ggpubr* (RRID:SCR_021139) were used for analysis and visualization. Categorical data were presented as percentages, whereas continuous data were expressed as means. The Mann-Whitney U test and Kruskal-Wallis test were used for comparisons between two and multiple groups, respectively. The Benjamini-Hochberg procedure was used to correct for multiple tests. The Kaplan–Meier method was used to visualize overall survival (OS), recurrence-free survival (RFS), recurrence-free probability (RFP), and distant control rate (DCR), and the log-rank test was used to test differences between groups. OS was defined as the time interval between the date of diagnosis and date of death. RFS was defined as the time interval between the date of diagnosis and date of recurrence or death. RFP was defined as the time interval between the date of diagnosis and date of recurrence. DCR was defined as the time interval between the date of diagnosis and date of distant recurrence. Cox proportional hazards survival regression was used to determine the influence of different variables on OS, RFS, RFP, and DCR. In the case of multiple significant associations in the univariate Cox regression analysis, the relevant variables were tested in a multivariate Cox regression analysis to identify independent prognostic factors. A *p*-value <.05 was considered statistically significant.

## Results

### Clinicopathological data

Overall, 54 patients with SGC were included in this study. The most frequent entity was salivary duct carcinoma (SDC; 43.6%), followed by acinic cell carcinoma (ACC; 24.1%), mucoepidermoid carcinoma (MEC; 22.2%), and secretory carcinoma (Sec; 11.1%). The mean age across the whole cohort was 57.7 (± 17.5) years, and 46.3% of all patients were female. Most patients (57.7%) had T1/2 tumors. Among SDC, 81.1% were androgen receptor (AR)-positive, and 54.5% showed nuclear AR positivity in >70% of tumor cells (AR_high_), whereas 45.5% showed nuclear AR positivity in ≤70% of tumor cells (AR_low_). Furthermore, 36.4% of the SDC were positive for human epidermal growth factor receptor 2 (HER2; score 3+ or HER2-amplified). The primary therapy was surgery in 98.1% of cases, and chemoradiation in 1.9% of cases. Neck dissection was performed in 74.1% of the cases, and 57.4% of the patients received adjuvant (chemo-)radiation therapy. Detailed data for the specific entities are presented in Table 1.

**Table 1.** Clinicopathological data.

|  | All<br>n = 54 (%) | SDC<br>n = 23 (%) | ACC<br>n = 13 (%) | MEC<br>n = 12 (%) | Sec<br>n = 6 (%) |
| --- | --- | --- | --- | --- | --- |
| Localization |  |  |  |  |  |
| Parotid gland | 50 (92.6) | 21 (91.3) | 13 (100.0) | 11 (91.7) | 5 (83.3) |
| Submandibular gland | 2 (3.7) | 1 (4.3) | 0 (0.0) | 0 (0.0) | 1 (16.7) |
| Small salivary gland | 2 (3.7) | 1 (4.3) | 0 (0.0) | 1 (8.3) | 0 (0.0) |
| Demographics |  |  |  |  |  |
| Female | 25 (46.3) | 5 (21.7) | 8 (61.5) | 9 (75.0) | 3 (50.0) |
| Male | 29 (53.7) | 18 (78.3) | 5 (38.5) | 3 (25.0) | 3 (50.0) |
| Age | 57.7 ± 17.5 | 66.5 ± 15.4 | 54.4 ± 18.3 | 48.8 ± 16.8 | 49.2 ± 18.8 |
| Histopathological parameters |  |  |  |  |  |
| T stage |  |  |  |  |  |
| T1/2 | 30 (57.7) | 11 (47.8) | 6 (50.0) | 9 (75.0) | 4 (80.0) |
| T3/4 | 22 (42.3) | 12 (52.2) | 6 (50.0) | 3 (25.0) | 1 (20.0) |
| N/A | 2 | 0 | 1 | 0 | 1 |
| N stage |  |  |  |  |  |
| N0 | 30 (57.7) | 5 (21.7) | 9 (75.0) | 11 (91.7) | 5 (100.0) |
| N+ | 22 (42.3) | 18 (78.3) | 3 (25.0) | 1 (8.3) | 0 (0.0) |
| N/A | 2 | 0 | 1 | 0 | 1 |
| M stage |  |  |  |  |  |
| M0 | 52 (96.3) | 21 (91.3) | 13 (100.0) | 12 (100.0) | 6 (100.0) |
| M1 | 2 (3.7) | 2 (8.7) | 0 (0.0) | 0 (0.0) | 0 (0.0) |
| Vascular invasion |  |  |  |  |  |
| V0 | 44 (81.1) | 17 (73.9) | 11 (91.7) | 11 (100.0) | 5 (100.0) |
| V1 | 6 (9.8) | 5 (26.1) | 1 (8.3) | 0 (0.0) | 0 (0.0) |
| N/A | 4 | 1 | 1 | 1 | 1 |
| Perineural invasion |  |  |  |  |  |
| Pn0 | 28 (56.0) | 4 (19.0) | 9 (75.0) | 10 (83.3) | 5 (100.0) |
| Pn1 | 22 (34.0) | 17 (81.0) | 3 (25.0) | 2 (16.7) | 0 (0.0) |
| N/A | 4 | 2 | 1 | 0 | 1 |
| Lymphovascular invasion |  |  |  |  |  |
| L0 | 37 (74.0) | 11 (50.0) | 10 (83.3) | 11 (100.0) | 5 (100.0) |
| L1 | 13 (26.0) | 11 (50.0) | 2 (16.7) | 0 (0.0) | 0 (0.0) |
| N/A | 4 | 1 | 1 | 1 | 1 |
| Extracapsular extension |  |  |  |  |  |
| ECE- | 41 (80.4) | 13 (59.1) | 11 (91.7) | 12 (100.0) | 5 (100.0) |
| ECE+ | 10 (19.6) | 9 (40.9) | 1 (8.3) | 0 (0.0) | 0 (0.0) |
| N/A | 3 | 1 | 1 | 0 | 1 |
| AR status (IHC) |  |  |  |  |  |
| Positive | N/A | 18 (81.1) | N/A | N/A | N/A |
| Negative | N/A | 4 (18.2) | N/A | N/A | N/A |
| N/A |  | 1 |  |  |  |
| >70% | N/A | 12 (54.5) | N/A | N/A | N/A |
| ≤ 70% | N/A | 10 (45.5) | N/A | N/A | N/A |
| N/A |  | 1 |  |  |  |
| HER 2 IHC Score |  |  |  |  |  |
| 0 | N/A | 5 (22.7) | N/A | N/A | N/A |
| 1+ | N/A | 4 (18.2) | N/A | N/A | N/A |
| 2+ | N/A | 5 (22.7) | N/A | N/A | N/A |
| 3+ | N/A | 8 (36.4) | N/A | N/A | N/A |
| N/A | N/A | 1 | N/A | N/A | N/A |
| Primary therapy |  |  |  |  |  |
| Surgical resection | 53 | 22 | 13 | 12 | 6 |
| Chemoradiation | 1 | 1 | 0 | 0 | 0 |
| Neck dissection |  |  |  |  |  |
| Yes | 40 | 21 | 11 | 10 | 4 |
| No | 12 | 1 | 2 | 2 | 2 |
| N/A | 2 | 1 | 0 | 0 | 0 |
| Adjuvant therapy |  |  |  |  |  |
| Radiation | 21 | 14 | 2 | 2 | 2 |
| Chemoradiation | 10 | 7 | 2 | 1 | 0 |
| None | 22 | 2 | 8 | 9 | 4 |
| N/A | 1 | 0 | 1 | 0 | 0 |

SDC: salivary duct carcinoma, ACC: acinic cell carcinoma, MEC: mucoepidermoid carcinoma, Sec: secretory carcinoma, AR: androgen receptor, HER2: human epidermal growth factor receptor 2, IHC: immunohistochemistry

With a median follow-up of 63.5 months for the entire cohort, SDC patients showed the most unfavorable five-year OS (50.6%), followed by MEC (83.3%), ACC (83.9%), and Sec patients (100.0%). Five-year RFS rates were 30.7% for SDC, 69.2% for ACC, 83.3% for MEC, and 66.7% for Sec. Moreover, five-year RFP was 39.9% for SDC, 69.2% for ACC, 91.7% for MEC, and 66.7% for Sec. Lastly, five-year DCR was 47.0% for SDC, 69.2% for ACC, 100.0% for MEC, and 83.3% for Sec.

### A 13-marker IMC panel identifies distinct immune cell and CAF subsets in nonMYO SGC

We used tissue from SGC primaries and metastases from 54 patients with SGC to create a tissue microarray (TMA). Tissue sections from TMA were stained with a cocktail of 13 metal-conjugated antibodies. Ablation of the tissue slices produced single-channel images that were used to generate multichannel images. After segmentation, the cells were categorized into cellular subsets. The cellular frequencies and results from the spatial analyses were then correlated with clinical parameters and patient outcomes (Figure 1A).

**Figure 1.**
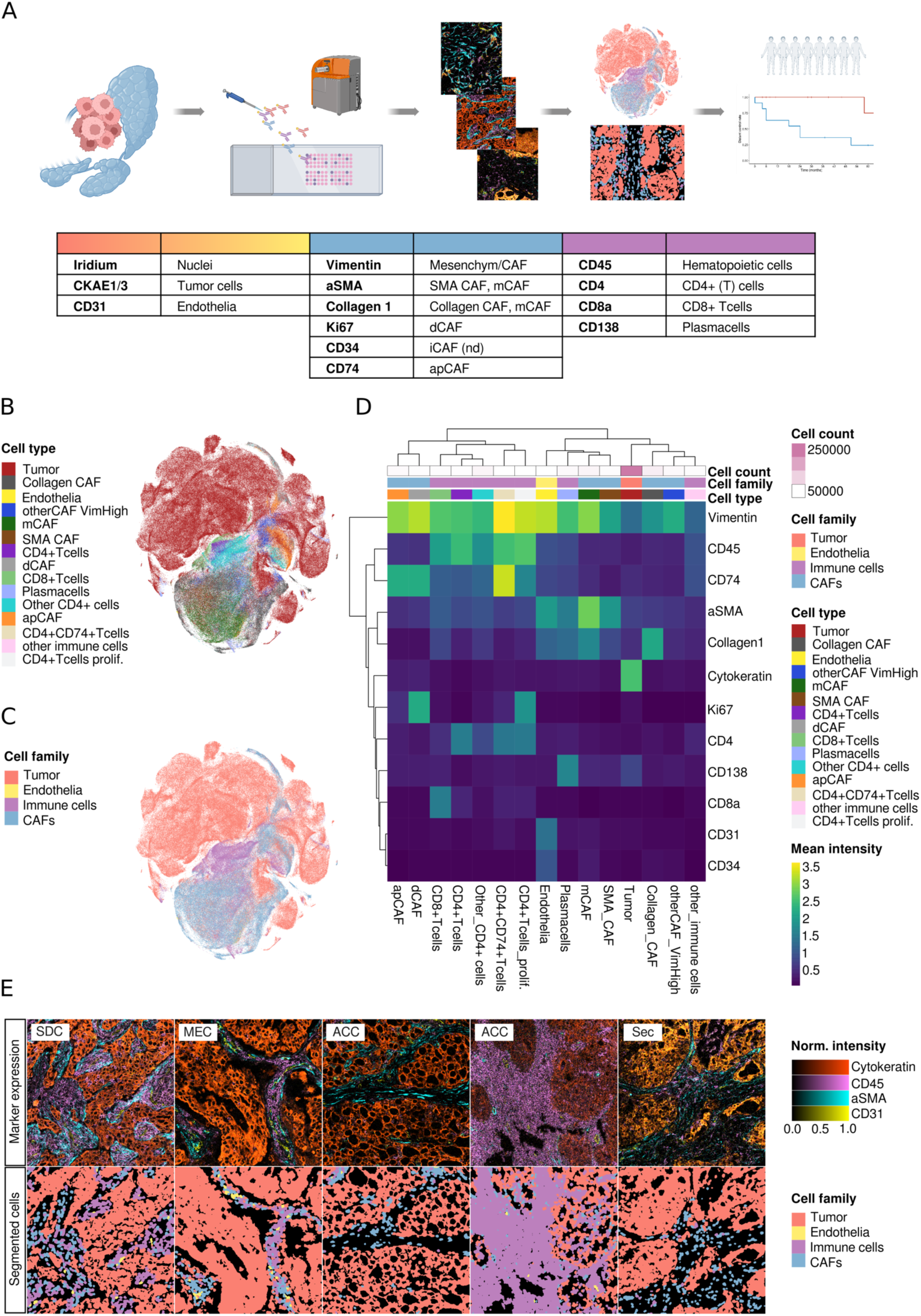
Phenotyping of the cellular SGC TME (A) Overview of the study: SGC samples from 56 patients were captured on a TMA, which was incubated with 13 metal-conjugated antibodies and ablated using a Hyperion system. Through image segmentation, a spatially-resolved single cell dataset was obtained and cell phenotyping was carried out in a semi-supervised fashion with established cell type markers. Cell frequencies and cellular neighborhoods were then correlated with clinical data. (B) tSNE plots of all 408.939 cells colored by cell type and (C) cell category. (D) Heatmap depicting the mean marker expression by cell type. (E) Representative images of the tumor architecture by marker expression (top row) and cellular composition after phenotyping (bottom row).

After the thorough exclusion of non-tumor-bearing TMA cores, we analyzed 199 SGC TMA cores from 54 SGC, including 188 primaries and 11 metastases from SDC, ACC, MEC, and Sec. A median cell count of 3,019 (ACC), 2,274 (Sec), 2,104 (MEC), and 2,029 (SDC) per 1 mm² ROI was noted. Using a stepwise Gaussian mixture model of cytokeratin expression and SOM clustering, we measured the expression of 13 different markers in 408,939 cells, we measured the expression of 13 different markers in 408,939 cells (Figure 1B and C). First, we separated tumor cells (CKAE1/3+; n = 266,172) from endothelial cells (CD31+, CKAE1/3−; n = 1,349), immune cells (CKAE1/3−, CD45+, or CD138+; n = 50,432), and CAFs (CKAE1/3/CD45/CD31/CD138−; n = 90,986). Immune cells were then clustered into plasma cells (CD138+), CD8+ T cells, CD4+ T cells, and weakly CD4-expressing cells, which are most likely monocytes or other myeloid-derived cells (other CD4+cells; (5, 12, 19)). CD4+ T cells were subclassified as CD4+CD74+ T cells, which most likely correspond to effector Tregs (20, 21), proliferating CD4+ T cells (Ki67+), and other CD4+ T cells. Immune cells solely expressing CD45 were classified as “other immune cells.” To classify CAFs, we adopted an IMC-validated classification scheme proposed by Cords et al. (5, 23), separating mCAFs (SMA+Collagen1+), Collagen CAFs (SMA-Collagen1+), SMA-CAFs (SMA+Collagen1-), dCAFs (ki67+, Vimentin+), and apCAFs (CD74+, Vimentin+). We also included CD73, IDO, and CD34 to detect rCAFs, ifnCAF/IDO CAFs, and iCAFs, respectively. However, the CD73 and IDO antibodies used did not produce a specific IMC signal. We could not discriminate iCAFs, since all CD34+ cells co-expressed CD31, a specific vascular marker, which was used to identify endothelia. Two subsets of cells did not display a specific marker profile: cells without expression of any of the markers used were designated as tumor cells, as they were located in tumor cell patches. Regarding the TME-centered focus of this work, we reasoned that this conservative approach would not impact TME-cell classification and thus minimize the chance of false-positive results. Cells with sole expression of Vimentin were classified as “other CAFs” with Vimentin expression (Figure 1D). Figure 1E displays representative ROIs of the included tumor entities as multiplex images and the corresponding cellular maps after segmentation and cell phenotyping.

### SGC entities are characterized by different TME cell frequencies

When pooling all ROIs, the cell type distribution across SGC tumor types displayed marked differences with a prominent immune cell compartment in ACC (Supplementary Figure S1C and D). No prominent differences in cell category contributions were noted when comparing primaries and metastases or peripheral and central tumor regions (Supplementary Figure S1E and F).

To account for differing numbers of ROIs per patient due to the exclusion of non-tumor-bearing and torn TMA cores, we calculated the median cell frequency per patient and compared the distribution of cells across the different SGC entities. We found a significantly varying abundance of immune cell subsets across tumor types (Figure 2A), which was largely due to significantly elevated levels of CD4+ T cells (p = 0.03), CD8+ T cells (p = 0.04), other CD4+ cells (p = 0.03), and unclassified immune cells (p = 0.04) in ACC. This resulted in a significantly higher frequency of overall immune cells in ACC (p = 0.04; Figure 2B and C).

**Figure 2.**
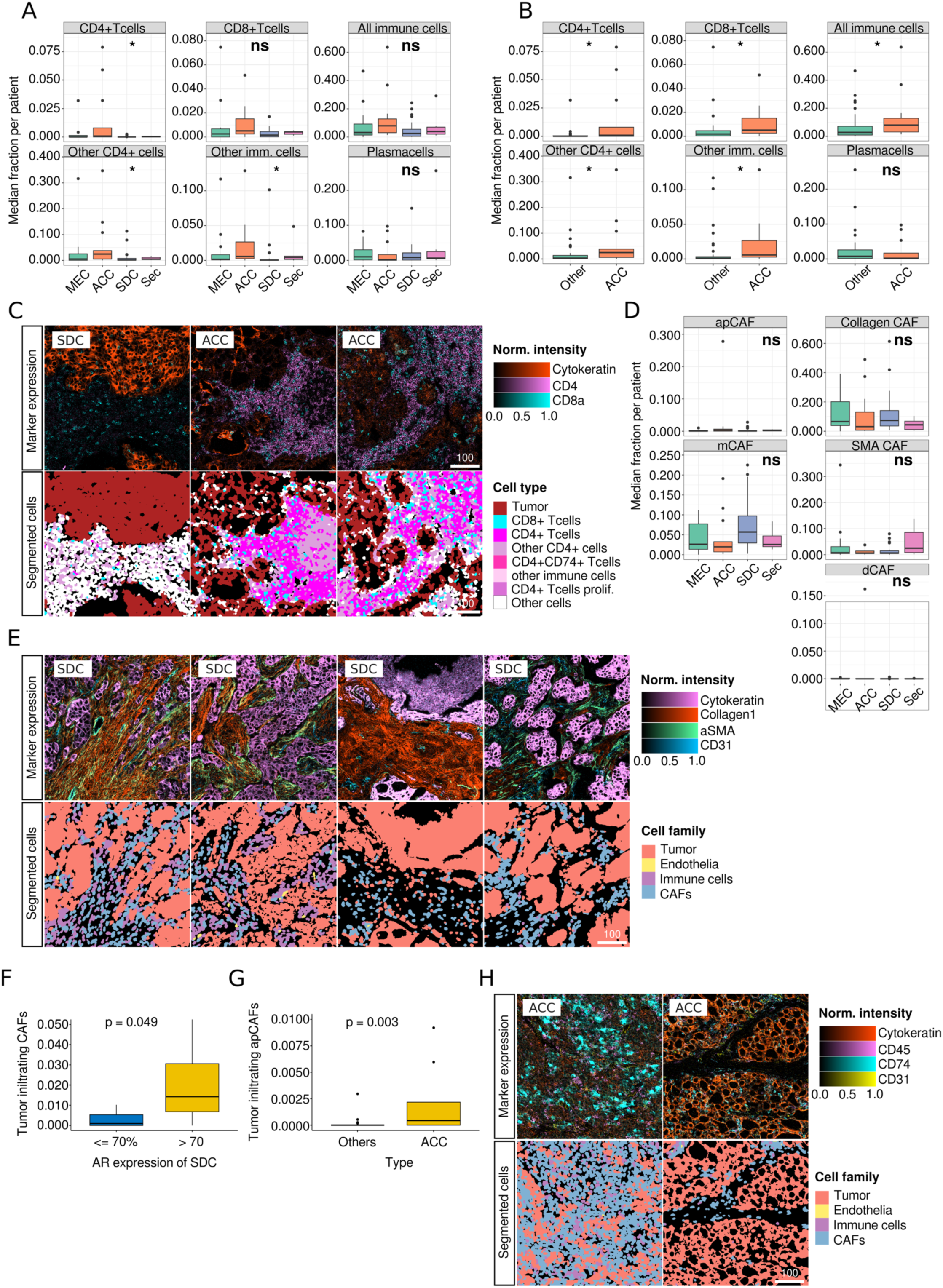
Comparison of median cell frequencies per patient across tumor subtypes (A) Immune cell subsets across SGC entities. (B) Immune cell subsets in ACC vs. other carcinoma types. (C) Representative core immune marker expression (top row) and selected immune cell subsets in SDC versus ACC. (D) Frequencies of CAF subtypes across SGC entities. (E) Examples of the spectrum of core CAF marker expression (top row) and cell categories (bottom row) in SDC. (F) Frequency of tumor-infiltrating CAFs dependent on AR expression in SDC. (G) Frequency of tumor-infiltrating apCAFs in ACC versus other SGC subtypes. (H) Representative images of CD74+ expression in apCAFs in ACC (top row) and corresponding cell categories after segmentation (bottom row).

Although the overall distribution of CAFs did not significantly vary in SGC types (Figure 2D), we detected the highest mean frequency of mCAFs in SDC, a CAF subset that has been associated with ECM production. However, in direct comparison with other entities, this elevation was not significant (Supplementary Figure S2A), probably because of the broad spectrum of mCAF frequencies within SDC (Figure 2D and E).

We also analyzed the frequency of tumor-infiltrating CAFs using tumor patch detection (Supplementary Figure S2B). As AR and HER2 are frequently expressed in SDC and are molecular targets for systemic therapy approaches (24), we examined the association between AR/HER2 expression and CAF frequency. We were able to show that the number of tumor-infiltrating CAFs was significantly elevated in AR_high_ SDC compared to AR_low_ tumors (p = 0.049; Figure 2F). No other significant cellular TME alterations were noted when SDC was subgrouped based on AR (Supplementary Figure S2C-E) and HER2 status (Supplementary Figure S2F-H). In contrast, in direct comparison to other tumor types, ACC displayed elevated levels of tumor-infiltrating apCAFs (p = 0.003; Figure 2G and H, Supplementary Figure S2I). However, the frequency of these cells was generally very low. No significant differences in tumor-infiltrating immune cells were noted across SGC tumor types (Supplementary Figure S2J).

### Spatial interaction analyses reveal a co-localisation of mCAFs with intratumoral vasculature

The spatial composition of the TME is an integral feature of tumor biology and influences the response to immunotherapy and patient outcomes (25). Therefore, we leveraged the spatial information preserved in the dataset.

After computing a spatial interaction graph (examples in Supplementary Figure S3A), we clustered cells based on their 20 nearest neighbors into 9 cellular neighborhoods (CN; Figure 3A and B). We found that the tumor cell-rich neighborhoods were largely devoid of CAFs and immune cells (CN1 and CN3). In contrast, CD4+ T cells, proliferating CD4+ T cells, other CD4+ cells, and CD8+ T cells formed a distinct lymphocyte-rich cluster (CN9, Figure 3C). Another immune-related CN mainly consisted of CD4+CD74+ T-cells, apCAFs, and dCAFs.

**Figure 3.**
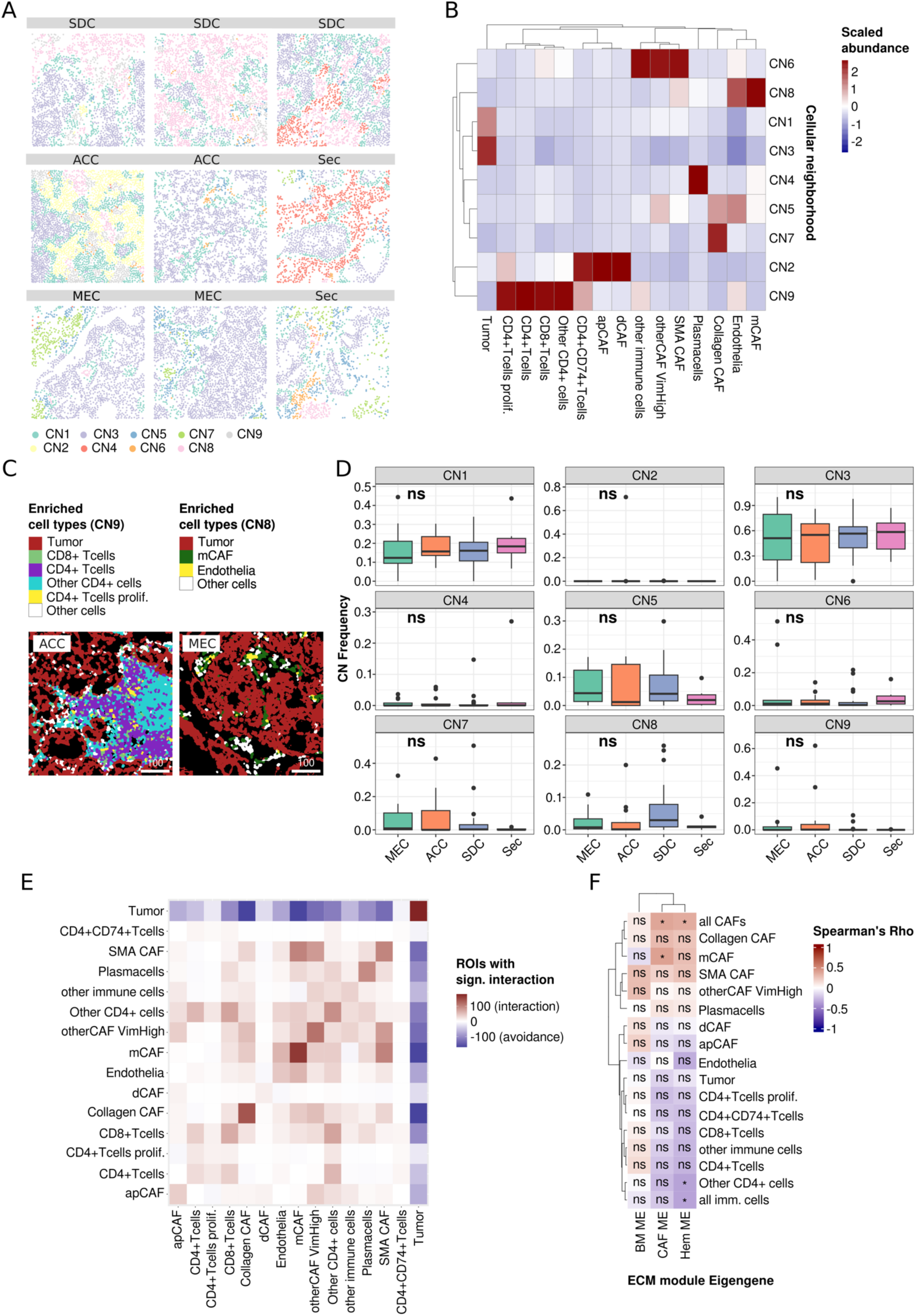
Spatial analysis of the TME of SGC and correlation with ECM protein modules (A) Exemplary ROIs with cells colored by cellular neighborhood (CN). (B) Scaled cell abundance per CN. (C) Representative ROI rich in cells that mainly contribute to CN9 (ACC) and CN8 (MEC). (D) Frequency of CN across SCG types. (E) Spatial interaction of cell types colored by the number of ROIs with significant co-localization of two cell types. A red color indicates more ROIs with significant interaction while blue tiles indicate more ROIs with significant avoidance of two cell types. (F) correlation of cell types with ECM module eigengenes which were published previously. Symbols within the tiles indicate the test significance (* p < 0.05, ns not significant).

However, as the contributing cell types, the latter CN occurred at very low frequencies (CN2, Supplementary Figure S3B). SMA CAF and mCAFs contributed to two distinct CN with partial overlap (CN 6 and 8, respectively; Supplementary Figure S3C). Interestingly, the matrix-associated CAF-rich (mCAF-rich) CN8 displayed a strong contribution to endothelia (Figure 3C). No specific marker for vCAFs, another cell type that was found to be associated with endothelia by Cords et al., was available in this panel. However, in contrast to mCAFs, very low expression of SMA and Collagen1 was reported in vCAFs (12), allowing for discrimination between the two cell types.

Although we did not observe significant differences in CN frequencies across SGC types, immune cell-rich CN was distinctively enriched in ACC. In addition, in SDC, we noted higher frequencies of the mCAF-rich CN8, but not of CNs 5, 6, and 7, which have high contributions of Collagen CAFs and SMA CAFs (Figure 3D, Supplementary Figure S3C and D).

The CN analyses were complemented by a more direct spatial interaction analysis, as proposed by Schapiro et al. (22) (Figure 3E). This approach compares the spatial interactions of cell type pairs with a null distribution in each ROI. The number of ROIs with significant positive (red) and negative (blue) interactions can then be summarized and graphically displayed. This methodically different analysis largely validated the aforementioned results, showing interactions among immune cells and between mCAFs and endothelia as well as SMA CAFs. Again, mutual exclusion of TME and tumor cells was noted.

In summary, we describe distinct spatial cellular TME neighborhoods in SGC that largely show a mutually exclusive predominance of tumor cells, immune cells, and CAFs. However, mCAFs and Collagen CAFs tend to localize in proximity to the tumor vasculature.

### CAFs and particularly mCAFs are associated with a distinct ECM profile

Recently, we dissected the ECM of SGC using an unbiased proteomic approach and discovered three protein clusters (“ECM modules”) that explain ECM differences across SGC tumor types via module *Eigengenes* (i.e., the resulting vector of the module’s proteomic signature) (15). Using gene set enrichment analysis, these modules were biologically annotated and found to be enriched for classic CAF-associated (CAF-module), basement membrane-associated (BM-module), and peripheral blood-associated (Hem-module) biological terms. The most significant members of the latter include coagulation-related factors such as kallikrein, kininogen, prothrombin, plasminogen, and angiotensin. Since CAFs are considered the main source of ECM in carcinomas, we leveraged an overlap of 40 patients between that and the present SGC cohort and correlated the module *Eigengenes* with the cell type frequencies (Figure 3F), thereby establishing a connection between the ECM and the cellular TME. As anticipated, we observed a strong positive correlation between CAFs with a more classic myofibroblastic phenotype (mCAF, Collagen CAF, SMA CAF) and the CAF protein module. However, only the correlation between mCAFs and overall CAFs was significant. In contrast, all immune cell subsets, except for plasma cells, were negatively correlated with the CAF module. Unexpectedly, very similar associations were found for the Hem-module, including an anti-correlation with endothelia and all immune cell subsets except plasma cells. We previously reasoned that this module consists of blood-related factors that are deposited within the ECM. However, the present data indicate that this does not imply exaggerated vasculature or deposition of cellular components of the peripheral blood. The BM module was mainly expressed in SGC with myoepithelial differentiation (e.g., adenoid cystic carcinomas), which were not analyzed in the present study. Together, these data clearly link SGC mCAFs, Collagen CAFs, and SMA CAFs to an increase in classical CAF-associated ECM proteins and provide evidence that the ECM components of the CAF and the Hem module participate in an immune-exclusive TME.

### A higher frequency of mCAFs is an independent prognostic factor for recurrence and distant metastasis in SDC

Cox regression analysis and log-rank test were performed to identify a potential influence of TME on the prognosis of SDC. The results of the univariate Cox regression model showed that a higher frequency of mCAFs was a prognostic factor for a higher rate of distant metastasis in SDC (p = 0.02; HR (95%CI) = 10.99 (1.37-88.16); Figure 4A). Regarding RFP, a higher frequency of mCAFs (p = 0.02; HR (95%CI) = 6.23 (1.34-28.93)) and a higher frequency of other CD4+ cells (p = 0.04; HR (95%CI) = 3.82 (1.07-13.69)) were prognostic factors for a higher probability of recurrence in the univariate Cox regression model (Figure 4B). Multivariate Cox regression revealed a higher frequency of mCAFs as an independent prognostic factor for a higher probability of recurrence (p = 0.025; HR (95%CI) = 5.9 (1.25-28.0); Figure 4C). The five-year DCR was 75.0% for patients with a low frequency of mCAFs and 24.2% for those with a high frequency of mCAFs (p = 0.0049; Figure 4D). T five-year RFP was 68.2% for patients with a low frequency and 18.2% for those with a high frequency of mCAFs (p = 0.0077; Figure 4E). A high frequency of CN8 (mCAF-rich) was marginally non-significant as a prognostic factor for a higher probability of recurrence (p = 0.06; HR (95%CI) = 4.54 (0.94-22.01); Supplementary Figure 4A) and distant metastasis (p = 0.07; HR (95%CI) = 3.46 (0.91-13.10); Supplementary Figure 4B) in the Cox regression model. Nevertheless, the five-year DCR was significantly lower in patients with a high frequency (27.7%) than in those with a low frequency of CN8 (68.2%; p = 0.04; Figure 4F). Further, the five-year RFP was 20.4% for patients with a high frequency and 61.4% for those with a low frequency of CN8 (p = 0.052; Figure 4G). Finally, a significantly higher frequency of mCAFs was found in patients with distant metastatic disease than in those without (p = 0.048). Sex did neither show significance as prognostic factor for DCR (p = 0.23; HR (95%CI) = 0.28 (0.03-2.25), nor for RFP (p = 0.36; HR (95%CI) = 0.48 (0.10-2.28). When including survival data, a higher frequency of mCAFs was not a prognostic factor for OS (p = 0.90; HR (95%CI) = 1.10 (0.34-3.63) or RFS (p = 0.16; HR (95%CI) = 2.10 (0.74-5.91) in univariate cox regression. Furthermore, the five-year OS did not differ significantly between patients with a high frequency of mCAFs (45.5%) and those with a low mCAF frequency (58.3%; p = 0.87). There was no difference in the five-year RFS between patients with a high mCAF frequency (18.2%) and those with a low mCAF frequency (43.7%; p = 0.15). Notably, patients with low mCAF frequencies were markedly older (mean age = 69.17 years) than those with high mCAF frequencies (mean age = 63.64 years).

**Figure 4.**
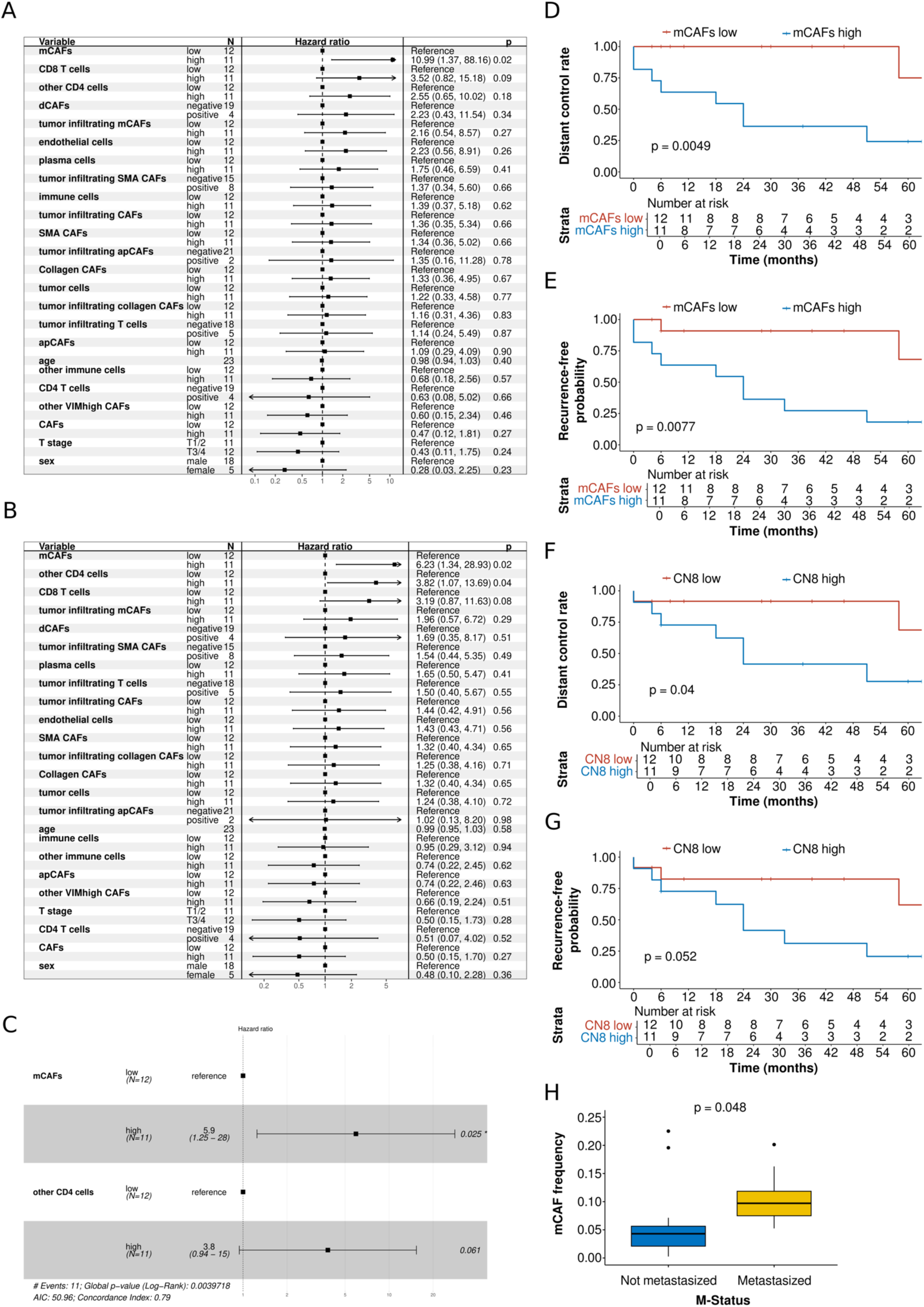
Association of cell types with distant control rate (DCR), recurrence-free probability (RFP), and frequency of distant metastasis in salivary duct carcinoma (SDC) patients (A) Univariate cox proportional hazards model for DCR for cell types and T stage, stratified by median proportion or as negative vs. positive in case median equals zero. N stage was excluded due to complete separation, resulting in an unbounded 95% confidence interval for its odds ratio. (B) Univariate cox proportional hazards model for RFP for cell types and T stage, stratified by median proportion or as negative vs. positive in case median equals zero. N stage was excluded due to complete separation, resulting in an unbounded 95% confidence interval for its odds ratio. (C) Multivariate cox proportional hazards model for RFP for mCAFs and other CD4 cells. (D) Kaplan-Meier plot with log-rank test for DCR comparing patients stratified as high and low based on the median proportion of mCAFs. (E) Kaplan-Meier plot with log-rank test for RFP comparing patients stratified as high and low based on the median proportion of mCAFs. (F) Kaplan-Meier plot with log-rank test for DCR comparing patients stratified as high and low based on the median proportion of CN8. (G) Kaplan-Meier plot with log-rank test for RFP comparing patients stratified as high and low based on the median proportion of CN8. (H) Boxplot displaying the proportion of mCAFs samples from patients with vs. without distant metastasis.

No prognostic tests were performed for entities other than SDC because of the low absolute number of events within those entities.

## Discussion

Although studies have described the tumor immune microenvironment (TIME) in a limited number of SGC entities (26, 27), there are no data on the composition of the non-immune cellular TME in SGC. The non-immune cellular TME mainly consists of tumor vasculature and CAFs. CAFs have been shown to directly influence tumor cells but also indirectly interact with them through the ECM and immune cells (28). Previous research has comprehensively demonstrated the pro-carcinogenic role of CAFs (29), but approaches specifically targeting CAFs have not been successful to date (13). Therefore, a more precise delineation of CAFs is necessary. Although different CAF subsets have been discovered in various solid tumor entities within the last years (8, 10, 30), a study by Cords et al. recently introduced a classification system for CAFs based on scRNA-seq data and demonstrated its applicability using IMC (5, 12). In the present study, we are the first to comprehensively decipher the cellular TME and evaluate the influence of cellular TME components on the prognosis of SGC.

We found that the highest mean frequency of mCAFs was observed in salivary duct carcinoma (SDC) samples compared to other entities. Importantly, in SDC, a higher mCAF frequency was an independent prognostic factor for a higher probability of recurrence and distant metastasis, and patients with a higher mCAF frequency had significantly worse DCR and RFP than those with a lower frequency of mCAFs. In the spatial interaction analysis, the mCAF-rich CN8 showed a strong contribution from endothelia. This is in line with previously published data that showed a positive spatial interaction of mCAFs with “blood,” high endothelial venules, and lymphatic channels in other solid tumors (5, 12). These findings suggest that mCAFs may play a role in facilitating hematogenous metastatic spread in SDC. A prior study has identified an anti-proportional distribution of mCAF and immune cell density in NSCLC and has hypothesized that mCAFs may negatively influence prognosis by shielding tumor cells from immune cells. However, our study is the first to suggest a potential role of mCAFs in hematogenous metastatic spread, possibly through the deposition of pro-tumorigenic factors such as ECM components, which have been shown to be associated with metastasis (31).

SDC is the clinically and biologically most aggressive SGC entity, with a five-year OS and RFS of approximately 50% and 30%, respectively, mainly due to distant recurrence (32–35). Although systemic therapy options targeting AR and HER2 are available for SDC, the median progression-free survival for drugs targeting both receptors is below nine months (36, 37). Therefore, new prognostic markers and therapeutic targets are urgently needed in this highly aggressive entity to guide treatment decisions in the curative setting and to improve outcomes in the R/M setting. Given the results of this study, the frequency of mCAFs could serve as a new prognostic marker for SDC. Dual immunohistochemical staining of SMA and Collagen1 could facilitate the quantification of mCAFs in FFPE samples at first diagnosis and thereby the stratification of patients into risk groups. Moreover, a targeted approach leading to a lower frequency of mCAFs in the primary tumor may decrease distant metastatic potential. To date, most clinical trials targeting CAFs have shown negative results (38). This is most likely because the relevant drugs were targeting CAFs in general rather than specific pro-tumorigenic subpopulations. For example, the FAP-targeting monoclonal antibody sibrotuzumab and the FAP-inhibiting small-molecule inhibitor talabostat did not show any objective response in phase II trials for metastatic colorectal cancer (39, 40). FAP has been found to be expressed in mCAFs, iCAFs, tCAFs, ifnCAFs, apCAFs, and dCAFs (among others) (5), some of which showed pro-tumorigenic and others anti-tumorigenic features (12). Identification of tumor-specific pro-tumorigenic subpopulations, such as mCAFs in SDC, and targeting these more specifically may help overcome the challenges faced in previous CAF-targeted therapies and lead to more effective treatment strategies.

Interestingly, we found a significantly higher frequency of tumor-infiltrating CAFs in AR_high_ than AR_low_ SDC cases. Although the impact of CAFs on tumor metabolism and signaling caused by cytokines and chemokines has been proven previously (41), there is currently no evidence showing the influence of CAFs on the AR status of tumor cells. While our data do not allow to draw causal conclusions in terms of the interplay between CAFs and AR expression in tumor cells, SDC patients with AR_high_ tumors could particularly benefit from CAF-targeted therapies or therapies combining androgen blockade and CAF-targeting.

With regard to the TIME, we found significant differences in the abundance of immune cell subsets across entities. This was due to higher levels of CD4+ T cells, CD8+ T cells, other CD4+ cells, and unclassified immune cells, leading to a significantly higher overall immune infiltration in ACC than in the other entities. The propensity for immune infiltration has been described as a characteristic of ACC in H&E diagnostics of these tumors (42); however, the exact cell types that contribute to this phenomenon have not yet been studied. Moreover, ACC has not been included in studies comprehensively describing the TIME in SGC (26, 27). Although ACC generally has a favorable prognosis, there is a subset of ACC patients with recurrence, resulting in a five-year RFS of approximately 76% (43) and requiring systemic therapeutic approaches. To date, there are no established therapeutic targets for ACC, and conventional chemotherapy has been shown to have low efficacy in SGC (44). Of note, programmed death (-ligand) 1 (PD(L)1)checkpoint blockade has not shown promising results in R/M SGC, with an objective response rate (ORR) of approximately 10% across entities (45, 46). This may be due to the relatively low expression of PD-L1 in SGC tumor cells, with a mean tumor proportion score (TPS) of approximately 4% across entities (47). According to our results, patients with ACC, compared to other SGC entities, could most likely benefit from new immunotherapeutic approaches that specifically target the TIME.

Furthermore, we found a strong positive correlation between the frequencies of mCAFs, SMA CAFs, and overall CAFs (classic myofibroblastic CAF phenotypes) and the CAF ECM protein module. This finding validates the previously suggested production of CAF ECM proteins by myofibroblastic CAF subpopulations (15). Additionally, all immune cell subsets, except for plasma cells, showed a negative correlation with the CAF module, which is consistent with the established immune-exclusive features of a stiffened carcinoma-associated ECM (48). Interestingly, the associations between the Hem module and cellular TME components, including an anti-correlation with endothelia, were very similar to those of the CAF module and its associations. We previously reasoned that the Hem module might consist of blood-related factors deposited within the ECM. However, the present data indicate that this does not imply exaggerated vasculature or deposition of the cellular components of the peripheral blood. The BM module had been shown to be mainly expressed in SGC with myoepithelial differentiation (e.g., adenoid cystic carcinomas) (15), which were not included in the present study. Together, these data clearly link SGC mCAFs, Collagen CAFs, and SMA CAFs to an increase in classical CAF-associated ECM proteins and provide evidence that the CAF and the Hem module are part of an immune-exclusive ECM.

This study has several limitations. First, SGG cases were identified retrospectively, which introduces the possibility of selection bias. Additionally, although relevant markers were included in the panel, we were unable to identify vCAFs, rCAFs, ifnCAFs, and iCAFs because of non-specific IMC signals or the unavailability of appropriate antibodies. Consequently, it is unclear whether these CAFs were not present in SGC or whether the absence of signals was due to the lack of specific metal-conjugated antibodies. Additionally, we did not include SGC types with (partial) myoepithelial differentiation, such as adenoid cystic carcinoma, because we identified a considerable overlap of myoepithelial and CAF marker expression during pretests, which would have resulted in high uncertainty in cell type identification. Lastly, the analyses were based on a limited number of cases and are therefore exploratory, which is a common problem in rare tumors, specifically in SGC showing a high number of biologically distinct entities.

This is the first study to comprehensively decipher the cellular TME in several of the more frequent SGC entities. To the best of our knowledge, this is the first study to use multiplexed imaging mass cytometry for spatial single-cell analyses in SGC. We were able to identify a high frequency of mCAFs as a prognostic factor for distant metastasis and further link mCAFs to hematogenous spread by spatial clustering in SDC. The frequency of mCAFs may be used to stratify patients with SDC into risk groups. Systemic therapy approaches targeting mCAFs in the TME of SDC could lead to improved prognosis in this highly aggressive tumor entity in the future. Moreover, we were able to demonstrate higher immune infiltration and cellular composition in ACC than in other SGC entities. Therefore, ACC may be amenable to new immunotherapeutic approaches. Overall, our results lay the groundwork for the improvement of diagnostics and therapy for these rare but clinically challenging carcinomas.

## Availability of data and material

The datasets generated and analyzed during the current study are available from the corresponding author upon reasonable request.

## Ethics approval

All procedures performed in this study were in accordance with the ethical standards of the institution or practice in which the studies were conducted (Approval Code: 13-091).

## Informed consent

Informed consent was obtained from all participants included in this study (no identifying information about the participants is available in this article).

## Consent for publication

All authors approved the final submitted manuscript.

## Data Availability

All data produced in the present study are available upon reasonable request to the authors

## Acknowledgements

The authors thank Petra Hofmann, Marion Müller, and Wiebke Jeske for their excellent technical support.

---

**Supplementary Figure S1:**
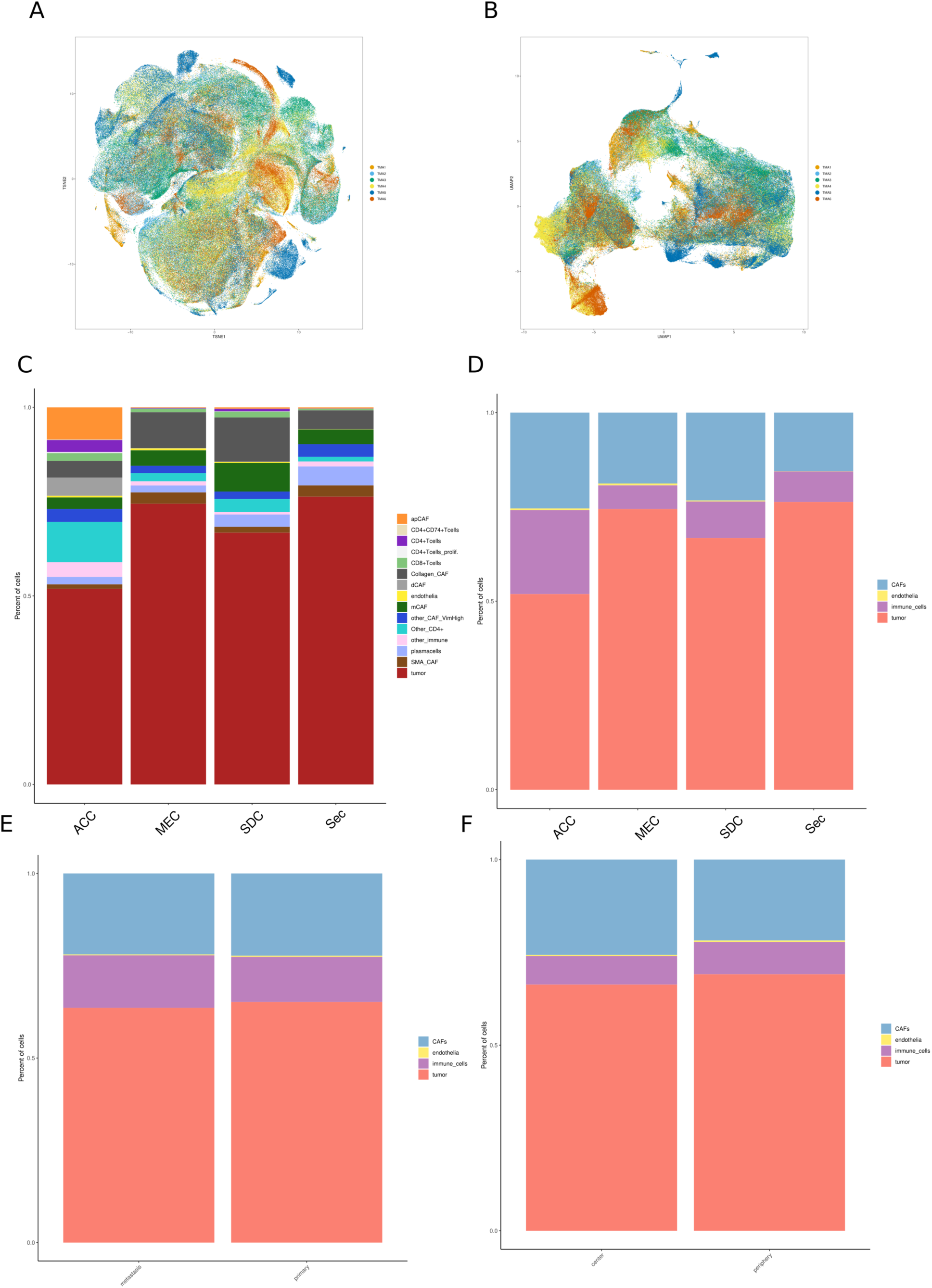
(A) tSNE and (B) UMAP projection of all analyzed cells colored by TMA. (C) Overall cell type and (D) cell category frequency. (E) Cell category frequency of primaries as well as metastases and (F) central and tumor compartments of the primary (all entities pooled).

**Supplementary Figure S2:**
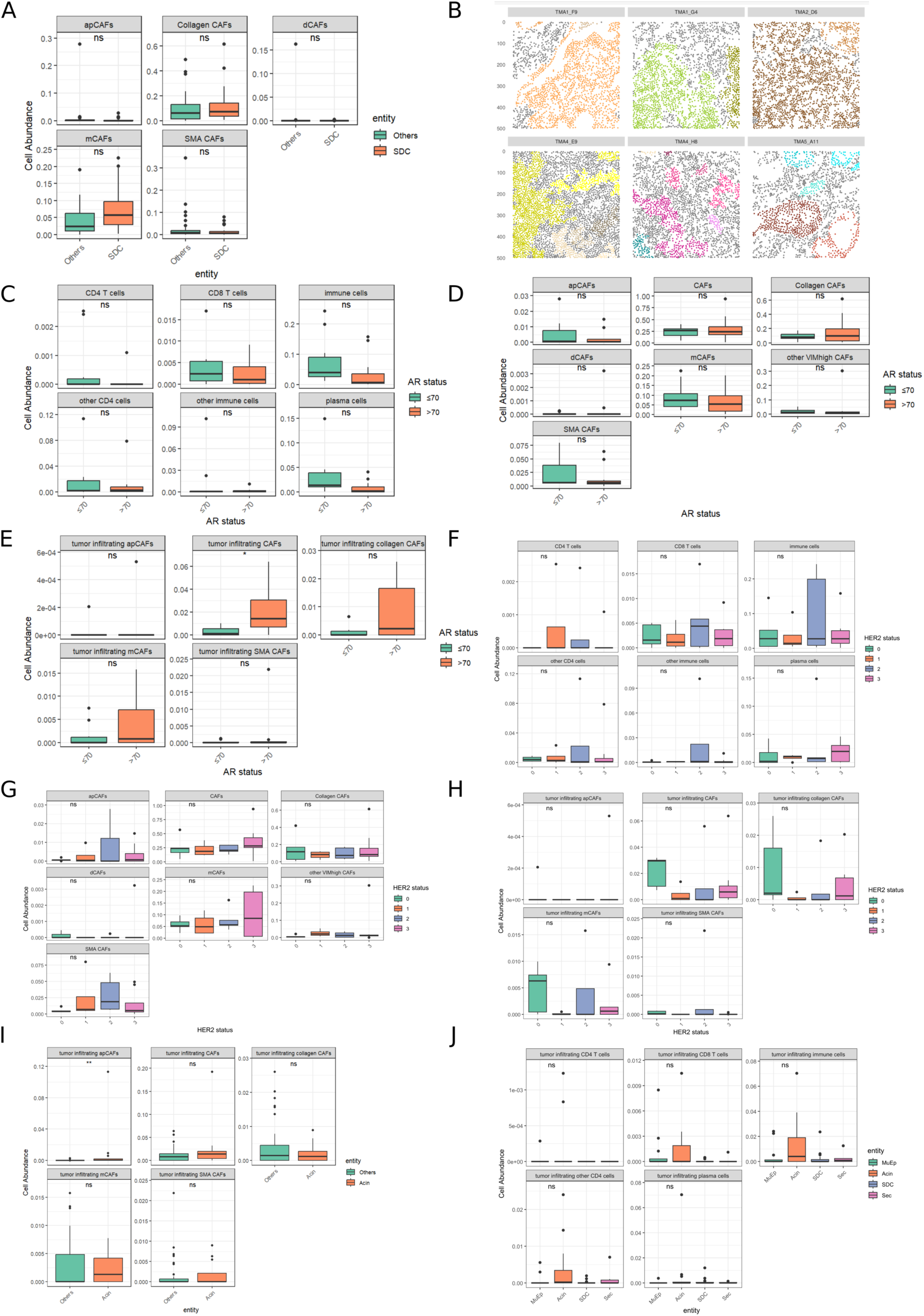
(A) Abundance of CAF subsets in SDC versus other tumor types. (B) Examples of tumor patch detection (tumor patches are colored, other cells depicted in grey). (C-E) Frequency of immune cells, CAFs and tumor-infiltrating CAFs compared between AR_high_ and AR_low_ SDC. (F-I) and between SDC with different HER2 expression levels. (J) Frequency of tumor-infiltrating immune cells in SGC entities.

**Supplementary Figure S3:**
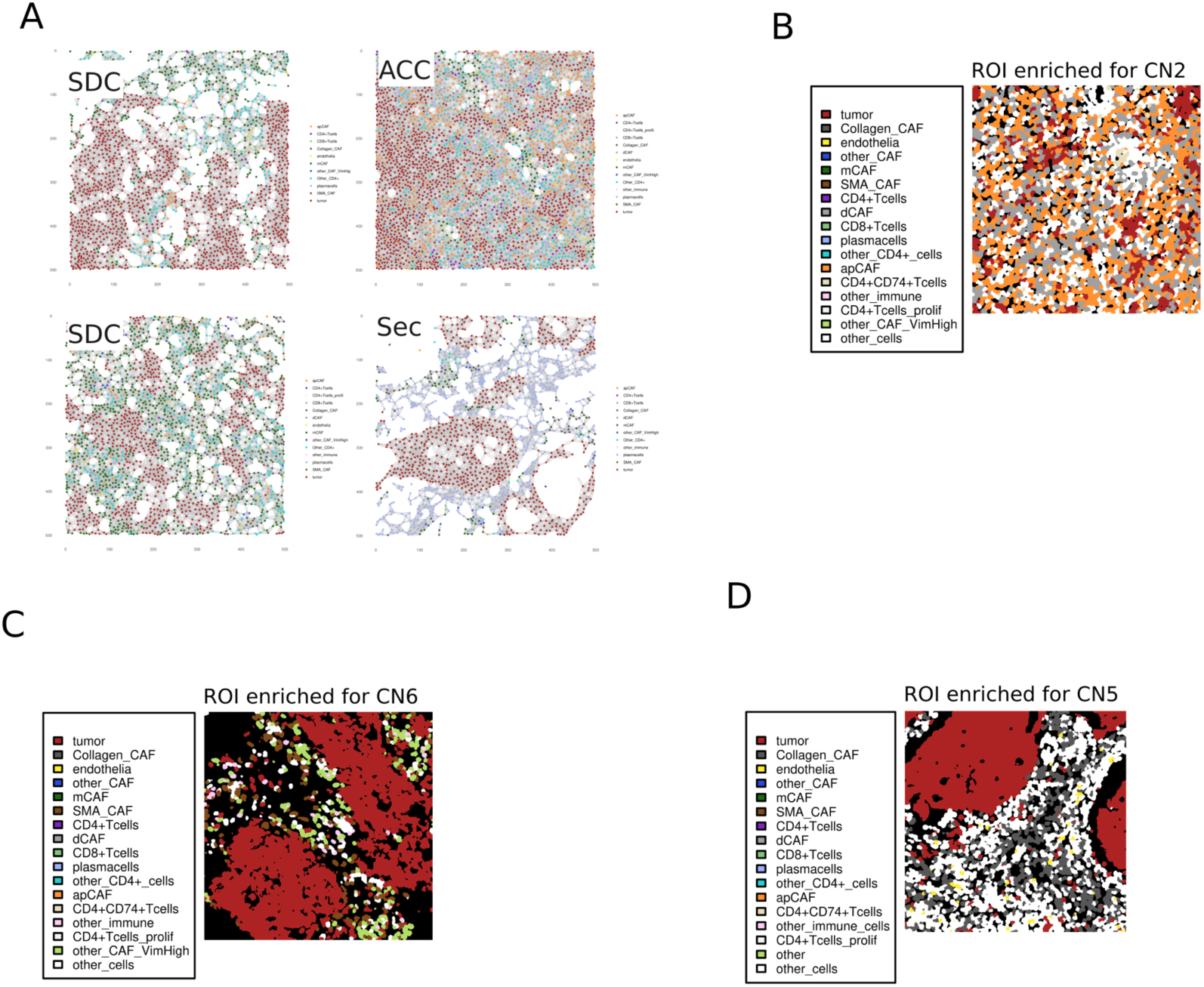
(A) Examples of cellular interaction graphs via k nearest neighbor detection with k = 20; colors correspond to cell types. (B-D) exemplary ROIs with co-localization of cells that are characteristic for cellular neighborhoods 2, 6, and 5.

**Supplementary Figure S4:**
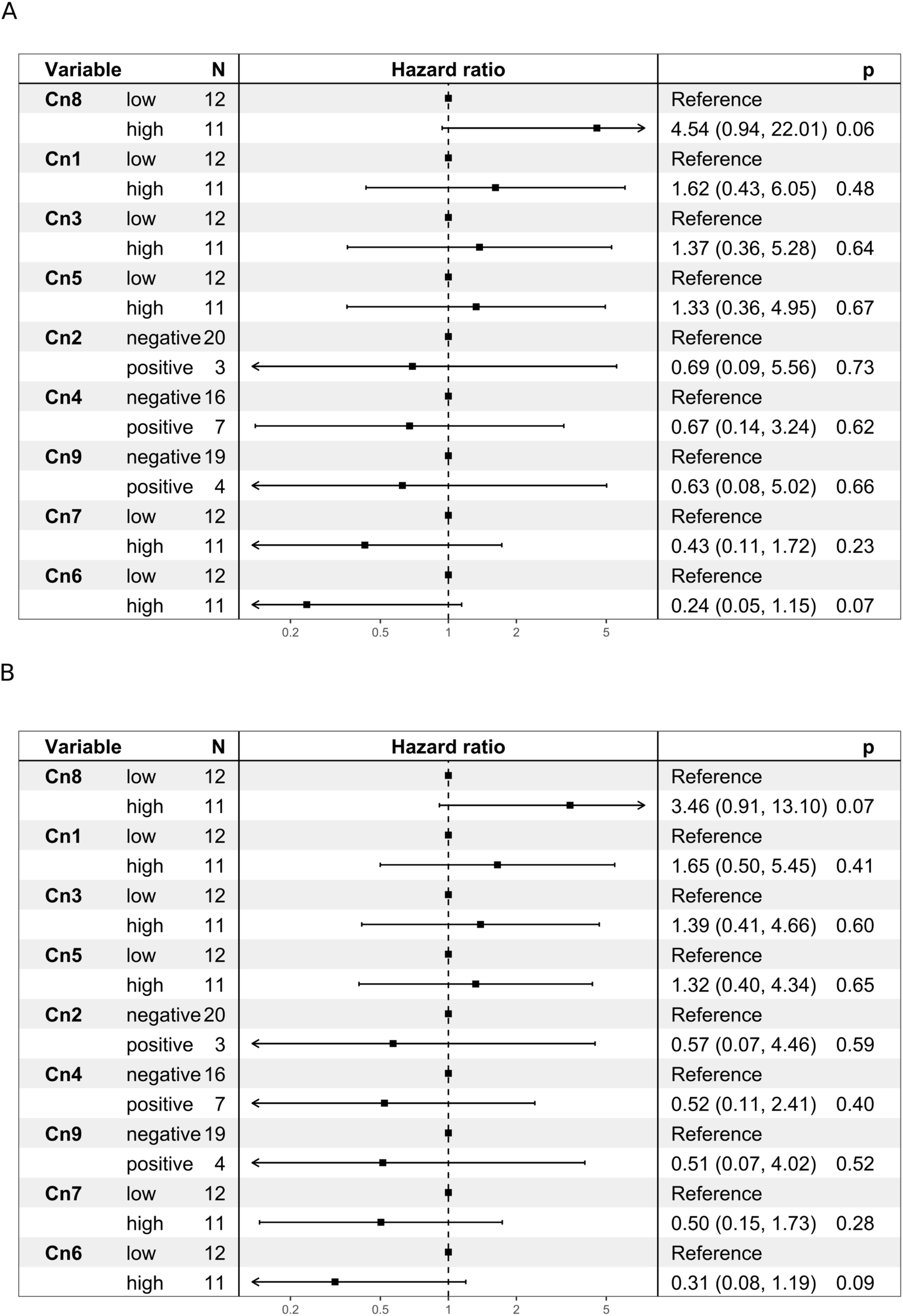
Univariate Cox proportional hazards model for (A) DCR and (B) RFP for cluster numbers (Cn) stratified by median proportion or as negative vs. positive in case median equals zero.

